# Global Research Architecture and Evolution of Neuroendoscopy for Intracranial Hemorrhage: A Bibliometric Analysis

**DOI:** 10.64898/2026.03.27.26349582

**Authors:** Zuoyue Duan, Mingyue Huang, Zhenghong Peng, Tianqi Tu

## Abstract

**Objective:** Neuroendoscopy has emerged as a crucial minimally invasive strategy for the treatment of intracranial hemorrhage (ICH). This bibliometric analysis aims to systematically delineate the global research architecture and evolution of neuroendoscopic ICH research over the past two decades.

**Methods:** Relevant publications were retrieved from the Web of Science Core Collection using a reproducible search strategy. Bibliometric tools were applied to analyze contributions from countries, institutions, authors, publications, keywords and journals, enabling the construction of a comprehensive knowledge map and evolutionary framework of this field.

**Results:** A total of 403 articles were identified, involving 2128 authors from 555 institutions across 43 countries. The publication trajectory exhibited fluctuating growth, reflecting the dynamic interplay between clinical demand and technological maturation. China contributed the highest publications and citation impact, followed by the US, jointly anchoring the global influence of the field. The research keywords have evolved from “intracerebral hemorrhage” and “initial conservative treatment” to “augmented reality.” Thematic evolution analysis revealed a clear progression from early emphasis on operative feasibility, safety, and perioperative outcomes toward more rigorous evidence appraisal and the refinement of context-specific clinical indications, accompanied by continuous technological innovation.

**Conclusion:** These findings collectively position neuroendoscopy as a cornerstone of modern ICH management, reshaping clinical strategies toward precision, minimal invasiveness, and multimodal intervention. Future progress will depend on strengthened international collaboration to generate high-quality evidence that supports patient stratification. The integration of emerging technologies, including advanced endoscopic robotics, is expected to further accelerate the translational and clinical landscape of neuroendoscopic ICH therapy.

## 1 Introduction

Intracranial hemorrhage (ICH) is a severe stroke subtype marked by bleeding within the cranium, leading to abrupt mass effect and a cascade of secondary injury [1]. Population-based studies indicate that the global incidence of ICH remains persistently high, ranging from approximately 19.1 to 26.5 per 100,000 person-years worldwide [2,3]. Moreover, ICH incidence increases markedly with advancing age [4], with a particularly pronounced elevation observed in individuals aged 65 years and older [5]. Early mortality remains strikingly high, with contemporary cohort data consistently reporting 30-day mortality rates of 30%–50% [6,7]. Survivors frequently experience severe long-term disability, functional independence at 90 days was achieved in only about 27% of patients [6].

Against the persistent unmet clinical need, especially in the elderly, surgical treatment for ICH was trapped for decades in skepticism. A succession of landmark randomized controlled trials (RCTs)—STICH [8], STICH II [9] and SWITCH [10]—fundamentally destabilized the credibility of traditional craniotomy, leaving the field urgently searching for new strategies. This impasse was disrupted by Scaggiante et al. [11], whose meta-analysis demonstrated reduced odds of death or dependence (odds ratio [OR] 0.40, 95% confidence interval [CI] 0.25–0.66) and mortality (OR 0.37, 95% CI 0.20–0.67) with neuroendoscopic surgery. These findings were reinforced by Hallenberger et al. [12], who reported higher 6-month favorable outcomes (p = 0.02) and lower mortality (p = 0.01). In this emerging paradigm, neuroendoscopy functioned as a conceptual shift of surgical intervention—reshaping the balance between therapeutic efficacy and iatrogenic injury cost, and signaling a cornerstone in the treatment of ICH.

Herein, we conducted the first bibliometric analysis to systematically portray the worldwide research landscape of neuroendoscopic treatment of ICH, highlighting thematic hotspots, evolutionary dynamics, and future directions, and offering a structured knowledge framework to guide future investigation.

## 2 Methods

### 2.1 Search strategy and data extraction

Literature related to neuroendoscopic treatment of ICH was retrieved from the Web of Science Core Collection (WOSCC). Articles published between January 1, 2006, and December 31, 2025 were identified using the following search strategy: TS=((neuroendoscop* OR “neuro-endoscop*” OR (“endoscopic surgery” NEAR/3 neurosurg*) OR (“endoscope-assisted” NEAR/3 (surgery OR evacuation)) OR ((endoscop* OR endoscope*) NEAR/3 (evacuat* OR aspirat* OR remov* OR “hematoma evacuation” OR “clot evacuation”))) AND (“intracranial hemorrhage” OR “intracerebral hemorrhage” OR “cerebral hemorrhage” OR “primary intracerebral hemorrhage” OR “spontaneous intracerebral hemorrhage” OR ICH OR “intraventricular hemorrhage” OR IVH OR “subarachnoid hemorrhage” OR SAH OR “subdural hematoma” OR “epidural hematoma”)). Only English-language publications were retained. Abstracts, reviews, conference proceedings, and editorials were excluded. All records were independently cross-checked by two researchers to remove unrelated publications. The final dataset was exported in tab-delimited format for subsequent bibliometric processing.

### 2.2 Data analysis and visualization

Bibliometric analysis was conducted using VOSviewer (version 1.6.20), Scimago Graphica (version 1.0.51, SCImago), Pajek (version 6.01) and CiteSpace (version 6.4.R2). Eligible records were imported into these platforms for network construction and visualization.

## 3 Results

### 3.1 General characteristics of publications

As shown in the workflow diagram (**Figure 1**), 403 studies were included in the final analysis. Between 2006 and 2025, 2128 authors from 555 institutions across 43 countries contributed to neuroendoscopic ICH research. The longitudinal publication profile revealed a non-linear yet overall ascending trend (**Figure 2A**), reflecting the gradual consolidation of academic interest in this area.

**Figure 1.**
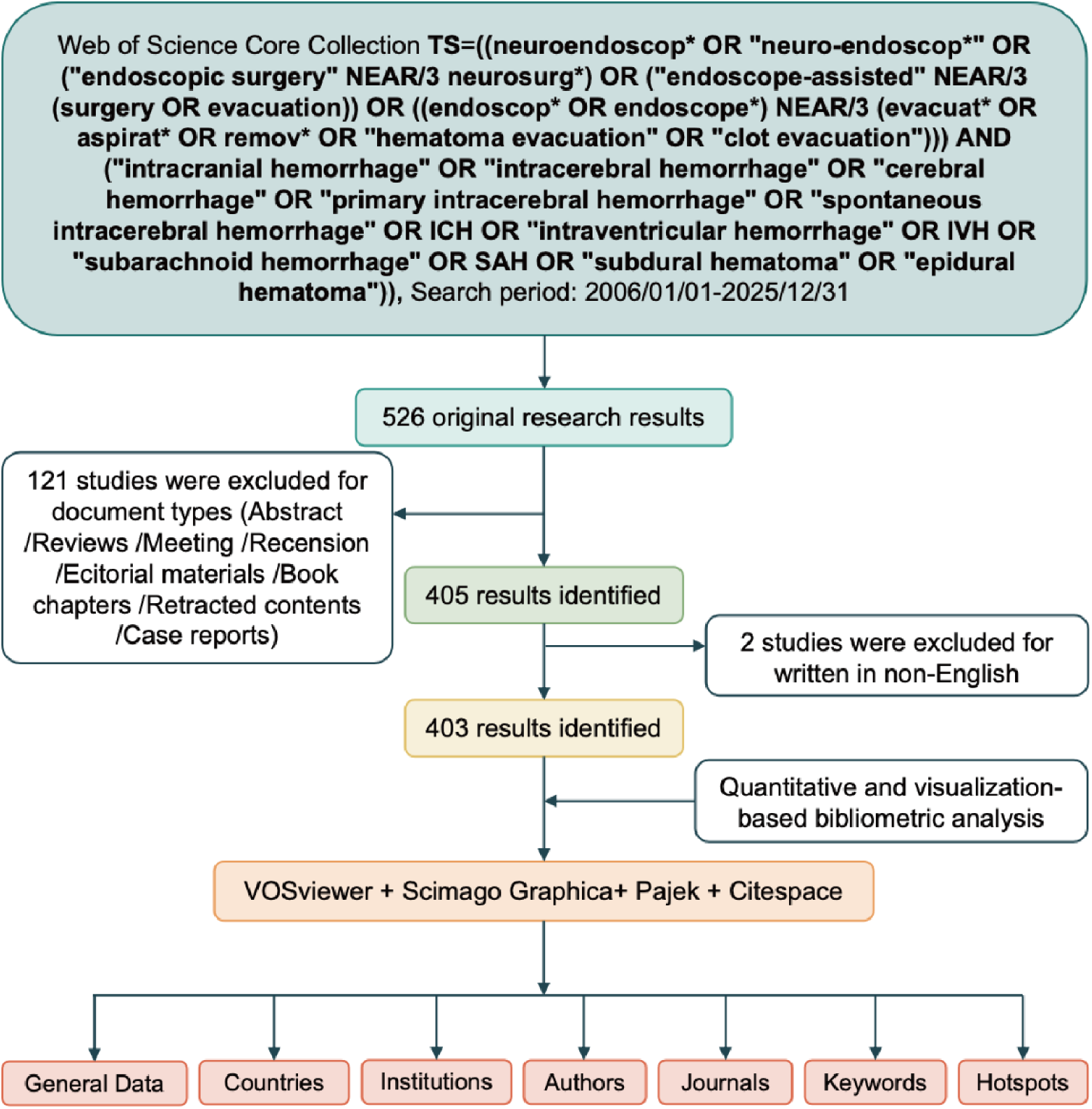
Literature selection strategy and analysis workflow.

**Figure 2.**
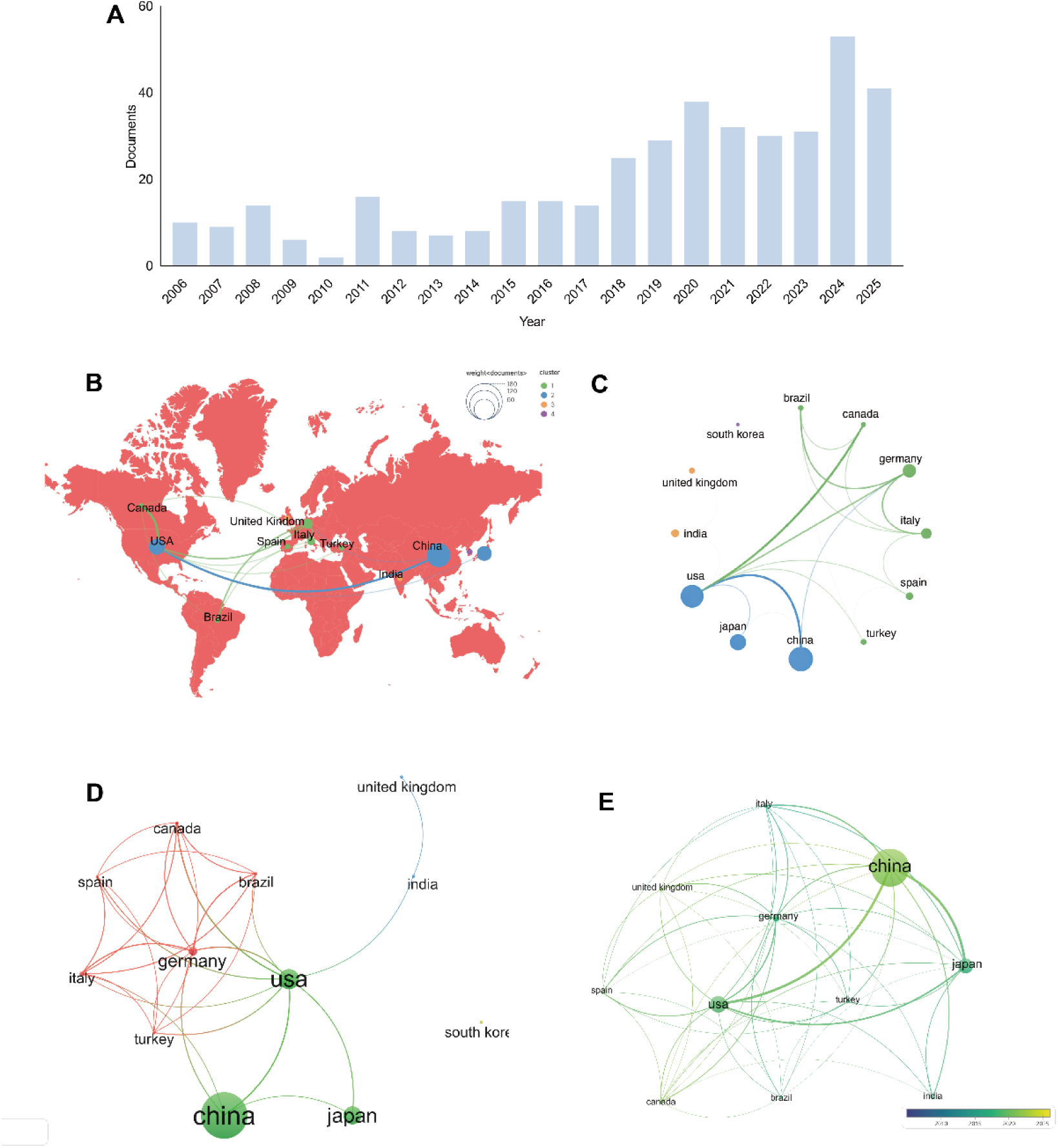
Country-level productivity, collaboration, and citation structure in neuroendoscopic ICH research. A, Annual publications. B, Geographical visualization of cooperative relationship between core countries, color-coded by clusters. Node size is proportional to the number of publications. Line thickness is proportional to co-authorship in the same article. C, The chord diagram of country cooperation. D, The cooperation networks. E, The citation network color-coded by publication timeline. Node size is proportional to the number of citations. Line thickness is proportional to link strength between nodes.

### 3.2 Country-level contribution and citation structure

Alongside the fluctuating rise in research attention, the country-level analysis further clarified the global distribution of contributions in this field. The 10 most active countries (**Figure 2B**) revealed a geographic concentration across Asia, North America, and Europe. China led with 168 publications (41.6%), followed by the US with 73 (18.1%). International collaboration patterns (**Figure 2B-2D**) indicated regional clustering with cross-regional co-authorship. The China–USA partnership was the central hub, connecting other active contributors, including Canada and European countries. Citation impact exhibited a parallel pattern (**Figure 2E**). China demonstrated the highest cumulative citation influence (1929), followed by the US (1647) and Japan (839). Overall, collaboration was led by China and the US, supporting the field’s evolution and providing a foundation for multicenter, cross-regional clinical studies.

### 3.3 Institution-level contribution and citation structure

Extending from the country-level analysis, we examined the 555 institutions (**Figure 3A**). The Icahn School of Medicine at Mount Sinai ranked first in publication volume (22), followed by Wuhan University (19) and Southern Medical University (10). The Icahn School of Medicine at Mount Sinai also exhibited the highest overall citation influence (534 total citations, 24.2 per paper). In contrast, the Chinese People’s Liberation Army General Hospital demonstrated the strongest average influence (192 citations, 32.0 per paper) despite fewer publications (6). These patterns reflected the institutions’ prominent scholarly engagement and sustained contributions to the development of the field.

**Figure 3.**
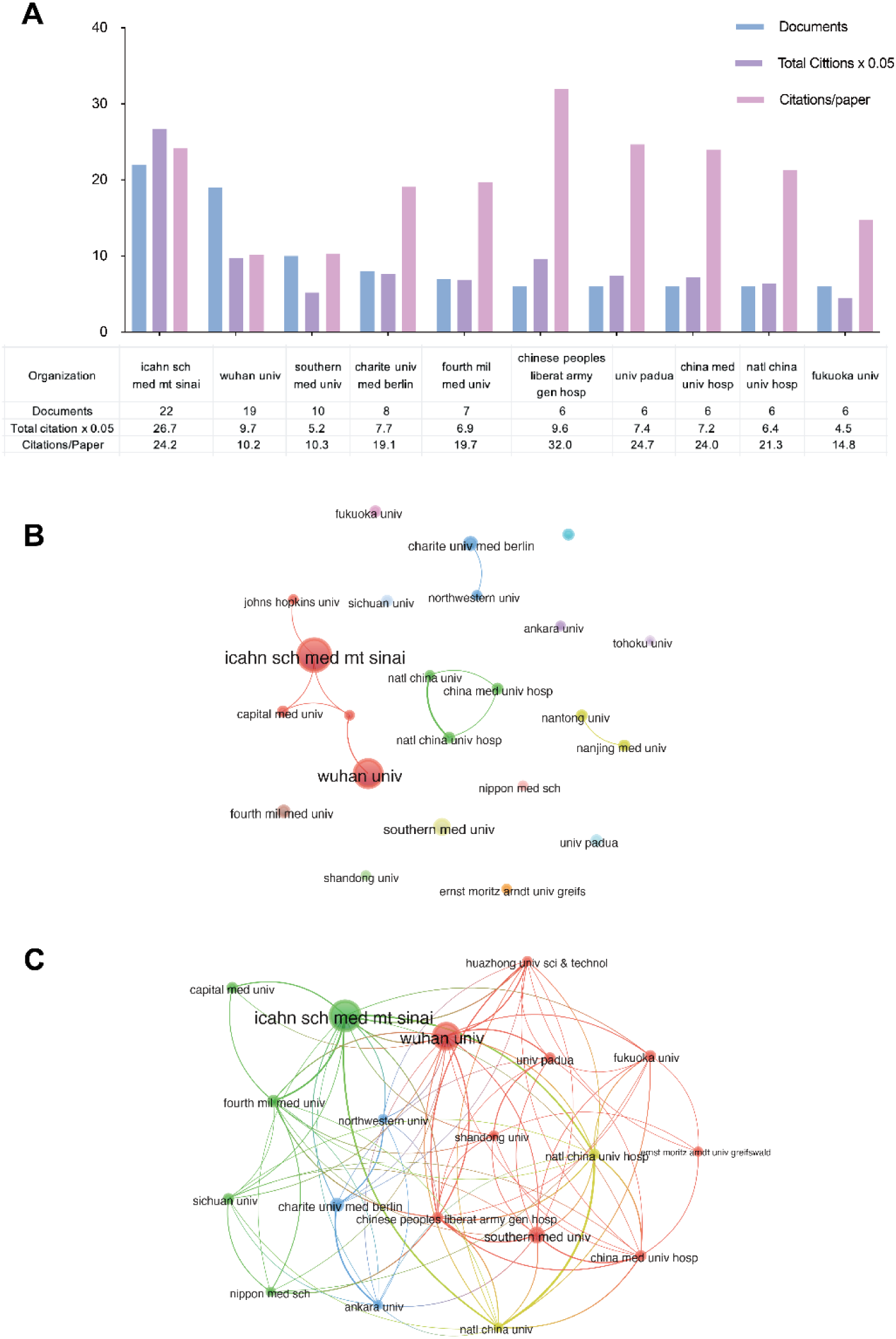
Institutional contributions and networks. A, Top 10 institutions with highest publications related to neuroendoscopic ICH. B, The cooperation network among institutions. C, The citation network among institutions.

Collaboration network analysis clarified the structural roles of key institutions (**Figure 3B**). The Icahn School of Medicine at Mount Sinai occupied a central collaborative position. Most leading institutions showed more independent research trajectories. The institutional citation network showed widespread reciprocal citations (**Figure 3C**). Wuhan University and the Icahn School of Medicine at Mount Sinai exhibited both high citation counts and stronger connectivity, indicating active knowledge circulation across centers. Overall, inter-institutional collaboration remained sparse, whereas cross-citation was active, highlighting substantial potential for deeper integrative partnerships.

Generally, core institutes have established a stable, influential presence, contributing high-volume and high-impact work. Future progress would benefit from major-center leadership to build coordinated platforms and reduce fragmented, uneven collaboration.

### 3.4 Author-level contribution and citation structure

At the author level, the collaboration network demonstrated a structured multi-cluster configuration (**Figure 4A**), forming research communities centered in the US, China, Germany, and Taiwan, aligning with the top 20 authors by publication volume (**Table 1**). Within both the US cluster (e.g., J. Mocco and C. P. Kellner) and the China cluster (e.g., Qiang Cai), central investigators exhibited dense and sustained collaborative linkages, reflecting stable, team-based structures. In contrast, inter-cluster connectivity remained relatively limited. Overall, this pattern showed strong regional cohesion but limited cross-regional integration, warranting multicenter collaboration.

**Figure 4.**
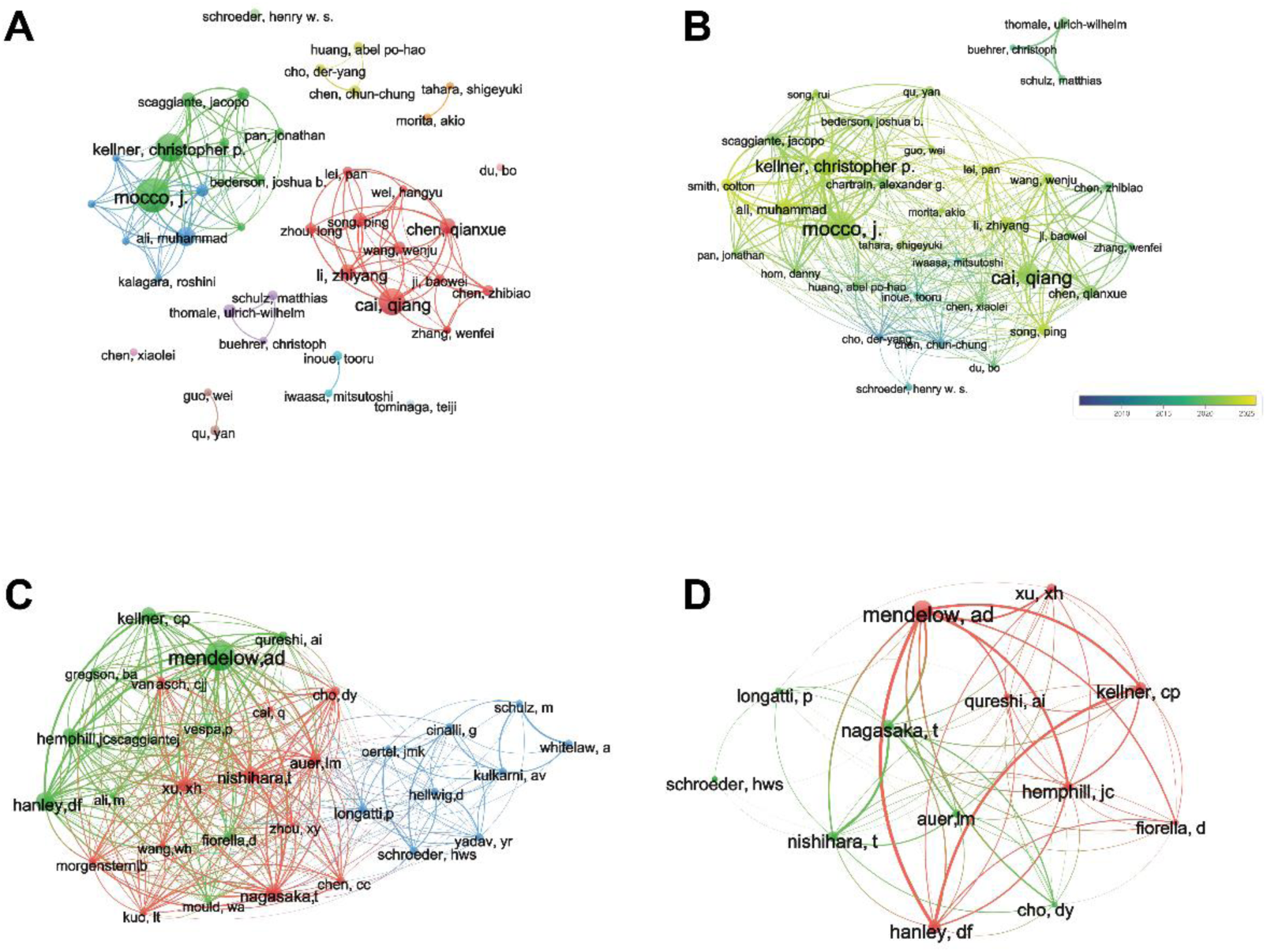
Author-level collaboration, citation structure, and thematic linking. A, The cooperation of authors. B, The citation network among authors color-coded by publication timeline. C, The co-citation network among authors with ≥30 citations. D, The co-citation network among authors with ≥50 citations.

**Table 1.** The top 20 authors with the highest publications on neuroendoscopic in intracranial hemorrhage research between 2006 and 2025.

| Rank | Author | Documents | Citations | Citations<br>/Paper | Country |
| --- | --- | --- | --- | --- | --- |
| 1 | Mocco, J. | 22 | 594 | 27.0 | US |
| 2 | Kellner, Christopher P. | 18 | 424 | 23.6 | US |
| 3 | Cai, Qiang | 17 | 190 | 11.2 | China |
| 4 | Ali, Muhammad | 12 | 137 | 11.4 | US |
| 5 | Chen, Qianxue | 10 | 101 | 10.1 | China |
| 6 | Li, Zhiyang | 10 | 75 | 7.5 | China |
| 7 | Scaggiante, Jacopo | 9 | 413 | 45.9 | Italy |
| 8 | Song, Ping | 9 | 79 | 8.8 | China |
| 9 | Chartrain, Alexander G. | 8 | 241 | 30.1 | US |
| 10 | Smith, Colton | 8 | 57 | 7.1 | US |
| 11 | Thomale, Ulrich-Wilhelm | 8 | 221 | 27.6 | Germany |
| 12 | Wang, Wenju | 8 | 71 | 8.9 | China |
| 13 | Bederson, Joshua B. | 7 | 245 | 35.0 | US |
| 14 | Chen, Chun-Chung | 7 | 309 | 44.1 | Taiwan,<br>China |
| 15 | Chen, Zhibiao | 7 | 83 | 11.9 | China |
| 16 | Zhou, Long | 7 | 45 | 6.4 | China |
| 17 | Cho, Der-Yang | 6 | 307 | 51.2 | Taiwan,<br>China |
| 18 | Hom, Danny | 6 | 197 | 32.8 | US |
| 19 | Huang, Abel Po-Hao | 6 | 128 | 21.3 | Taiwan,<br>China |
| 20 | Inoue, Tooru | 6 | 89 | 14.8 | Japan |

Citation network and timeline analyses delineated three phases (**Figure 4B**). In 2005–2015, researchers primarily from Taiwan and mainland China (notably Der-Yang Cho and Chun-Chung Chen) established technical feasibility and procedural safety [13–16]. In 2015–2020, German groups extended applications to pediatric post-ICH hydrocephalus [17–21]. since 2020, China and the US have led with complementary emphasis—technique iteration in China [22–25] and evidence-based indication and outcome refinement in the US [26–30]. Consistently, co-citation analysis demonstrated a regionally structured knowledge architecture (**Figure 4C–D**). Using a ≥30-citation cutoff (**Figure 4C**), three communities—North America, East Asia, and Europe—emerged. The North American cluster, anchored by Mendelow AD [8,9] and Hanley DF [31] retained centrality at ≥50 citations (**Figure 4D**), underscoring its foundational evaluative role. The East Asian cluster emphasized comparative analyses of surgical strategies [32,33], technical refinement [34], and clinical indications [33] and persisted at ≥50 citations (**Figure 4D**). The European cluster focused on specialized domains such as intraventricular hemorrhage [35] and post-ICH hydrocephalus [21], which were less integrated and did not remain at ≥50 citations (**Figure 4D**). Collectively, these patterns indicated that neuroendoscopic ICH research has matured into a distinct research domain characterized by a complementary intellectual structure that integrates North American evidence-oriented paradigms, East Asian technical and instrumental innovation, and European domain-specific thematic expansion.

The three-field plot (**Figure 5A**) showed a temporal shift from general ICH management and craniotomy comparisons to minimally invasive and endoscopic paradigms, with tighter integration with specific clinical contexts, including hypertensive ICH and ICH complicated by intraventricular hemorrhage and hydrocephalus. Citation burst analysis (**Figure 5B**) suggested a relay-like evolution of priorities rather than clear-cut temporal discontinuities. From 2017–2020, Scaggiante, Zhibiao Chen, Bo Du, and Qianxue Chen emphasized technical legitimacy, with attention on single-port access and operative trajectories [22,36,37]. from 2020–2022, Kellner, Hangyu Wei, and Rui Song shifted to comparative minimally invasive surgery (MIS) evaluations, focusing on early outcomes with mixed conclusions [26,28]. Overall, the relay-like pattern highlights agenda inheritance and translation from technical validation to clinical positioning.

**Figure 5.**
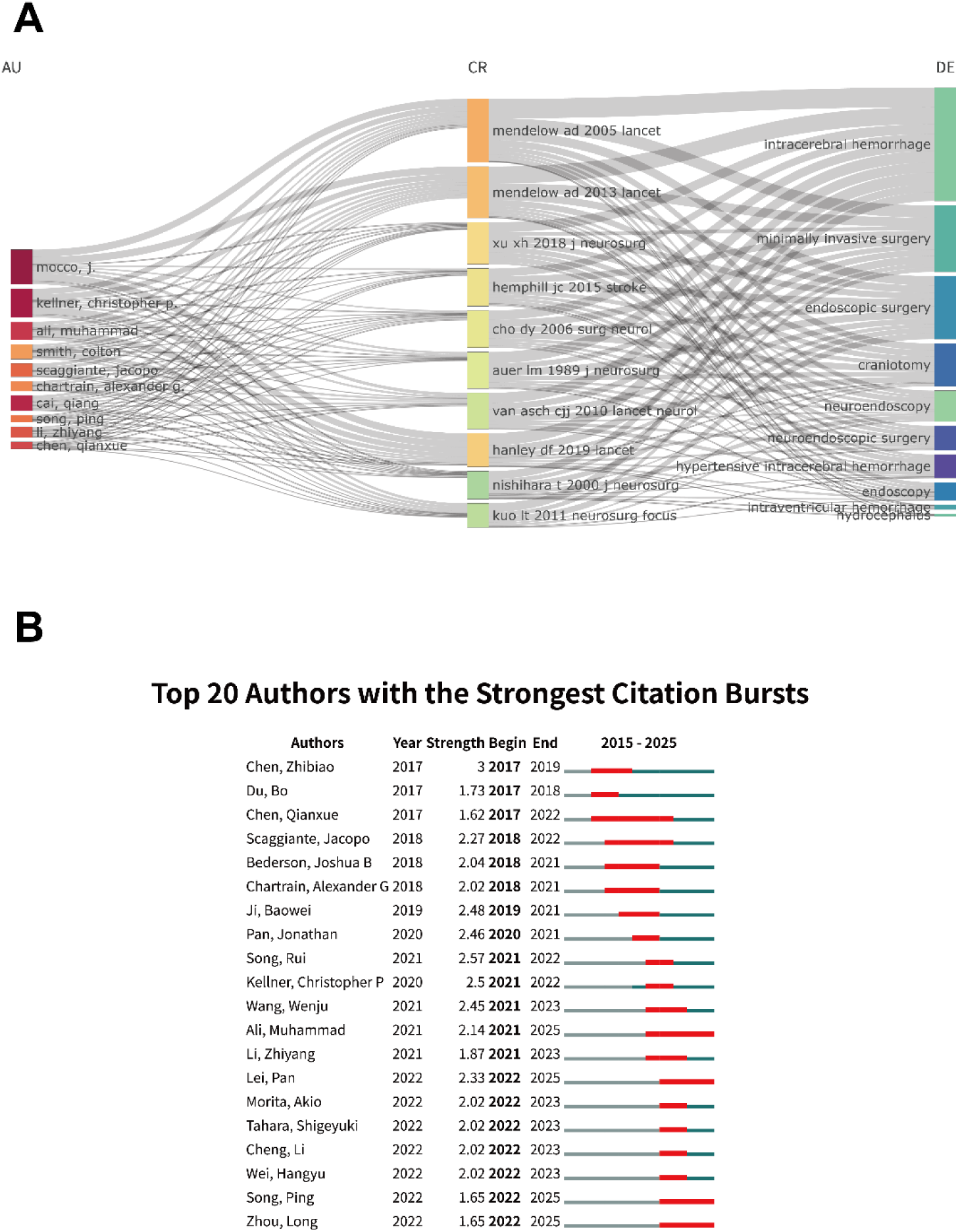
Keywords Plus analysis and citation bursts of authors in neuroendoscopic ICH research. A, Three-field plot of the Keywords Plus analysis on neuroendoscopic ICH (Left field: authors; Middle field: cited references; Right field: descriptive keywords). Bjl, Top 20 authors with the strongest citation bursts. Red bars indicate high citations in the year.

Collectively, neuroendoscopic ICH research evolved from feasibility to evidence-based evaluation, refined stratification, and long-term outcome assessment, supporting next steps in mechanistic work, multicenter collaboration, standardization, and guideline integration.

### 3.5 Publication-level contribution and citation structure

At the publication level, the most highly cited articles were summarized in **Table 2**. The citation network (**Figure 6A**) revealed sparse co-citation concentrated in a few core works. The most cited study was Scaggiante et al. (Stroke, 2018; 175 citations) [37], a meta-analysis of 15 RCTs (1989–2016) comparing MIS with conservative care and conventional craniotomy. MIS reduced death or dependence (OR 0.46, 95% CI 0.36–0.57) and mortality (OR 0.59, 95% CI 0.45–0.76), with endoscopic surgery showing similar associations (death or dependence OR 0.40, 95% CI 0.25–0.66; mortality OR 0.37, 95% CI 0.20–0.67). Beyond effect estimates, it helped re-anchor the field toward MIS after trials such as STICH II [9] tempered confidence in routine craniotomy.

**Figure 6.**
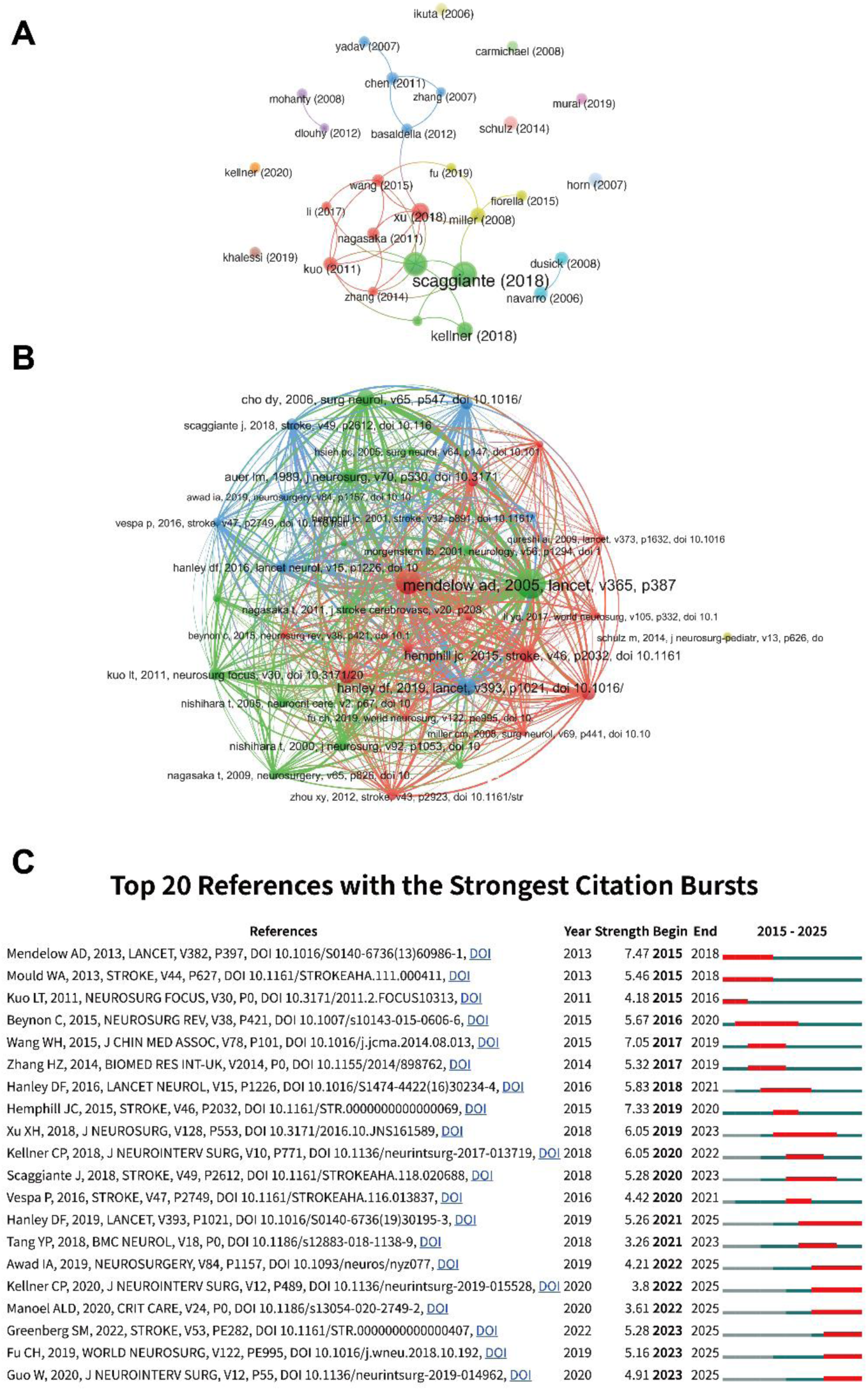
Document-level citation structure and burst detection. A, The citation network among eligible articles. B, The co-citation network among publications. C, Top 20 references with the strongest citation bursts.

**Table 2.** The top 10 references with the highest citations on neuroendoscopic in intracranial hemorrhage research between 2006 and 2025.

| Rank | Author | Title | Journal | Year | Citations |
| --- | --- | --- | --- | --- | --- |
| 1 | Scaggiante, J | Minimally Invasive Surgery for Intracerebral Hemorrhage: An Updated Meta-Analysis of Randomized Controlled Trials | Stroke | 2018 | 175 |
| 2 | Cho, DY | Endoscopic surgery for spontaneous basal ganglia hemorrhage: comparing endoscopic surgery, stereotactic aspiration, and craniotomy in noncomatose patients | Surgical Neurology | 2006 | 165 |
| 3 | Xu, XH | Effectiveness of endoscopic surgery for supratentorial hypertensive intracerebral hemorrhage: a comparison with craniotomy | Journal of Neurosurgery | 2018 | 116 |
| 4 | Kellner, CP | The Stereotactic Intracerebral Hemorrhage Underwater Blood Aspiration (SCUBA) technique for minimally invasive endoscopic intracerebral hemorrhage evacuation | Journal of Neurointerventional surgery | 2018 | 104 |
| 5 | Miller, CM | Image-guided endoscopic evacuation of spontaneous intracerebral hemorrhage | Surgical Neurology | 2008 | 94 |
| 6 | Kuo, LT | Early endoscope-assisted hematoma evacuation in patients with supratentorial intracerebral hemorrhage: case selection, surgical technique, and long-term results | Neurosurgical Focus | 2011 | 88 |
| 7 | Schulz, M | Neuroendoscopic lavage for the treatment of intraventricular hemorrhage and hydrocephalus in neonates Clinical article | Journal of Neurosurgery-Pediatrics | 2014 | 86 |
| 8 | Horn, EM | Treatment options for third ventricular colloid cysts: Comparison of open microsurgical versus endoscopic resection | Neurosurgery | 2007 | 86 |
| 9 | Navarro, R | Endoscopic third ventriculostomy in children: early and late complications and their avoidance | Childs Nervous System | 2006 | 83 |
| 10 | Dusick, JR | Success and complication rates of endoscopic third ventriculostomy for adult hydrocephalus: a series of 108 patients | Surgical Neurology | 2008 | 80 |

The co-citation network (**Figure 6B**) showed a knowledge backbone anchored by three landmark studies STICH (2003) [38], which challenged routine craniotomy in ICH; the 2015 guideline statement [39], which framed evidence-based management and key gaps; and MISTIE III (2019) [31], which reappraised MIS, showing no significant improvement in favorable outcomes versus medical care (p = 0.33) but fewer 30-day serious adverse events (p = 0.012). Subsequent neuroendoscopic and strategy-focused studies were tightly linked to these anchors, indicating iterative refinement around this core.

Reference citation burst analysis (**Figure 6C**) extended from landmark RCTs (STICH II [9]; MISTIE III [31]) and consensus guidelines [39] strategy-oriented syntheses (e.g., the meta-analysis by Scaggiante et al. [11]). Earlier bursts centered on pivotal RCTs on traditional craniotomy, whereas recent bursts aligned with neuroendoscopic clinical studies refining procedural indications and application scenarios [40]. Collectively, this trend indicated a shift from foundational evidence to context-specific, technique-oriented clinical exploration.

Overall, publication-level analysis indicates that neuroendoscopic ICH research evolved by consolidating and reinterpreting core evidence base anchored by RCTs, guidelines, and meta-analyses. These anchor works served as conceptual and methodological reference points that shaped study design and clinical interpretation, and gradually shifted the field from foundational validation to indication- and scenario-specific refinement.

### 3.6 Keyword occurrence and co-occurrence analysis

Building on the publication-based analysis, keyword-level analysis further clarified the field’s evolving thematic focus over the past two decades. Accordingly, Keywords Plus was used instead of author-provided keywords. As an algorithmically derived indexing system based on cited references, Keywords Plus captures broader conceptual linkages and improves sensitivity in identifying latent thematic structures in bibliometric mapping.

The Keywords Plus co-occurrence network (**Figure 7A**) was structured around three principal dimensions: operative intervention, comparative therapeutic strategies, and clinical application contexts. Core terms such as “management” and “surgery” occupied central positions, indicating that clinical decision-making and operative management consistently constituted the conceptual nucleus of the field. Surrounding this core, comparative therapeutic concepts, including “early surgery” and “initial conservative treatment”, formed a distinct analytical cluster. In parallel, broader clinical contexts such as “stroke” and “spontaneous ICH” provided the overarching framework of application. Taken together, this configuration reflected the consolidation of neuroendoscopic ICH research into an integrated conceptual structure linking decision processes, procedural strategies, and comparative evidence assessment.

**Figure 7.**
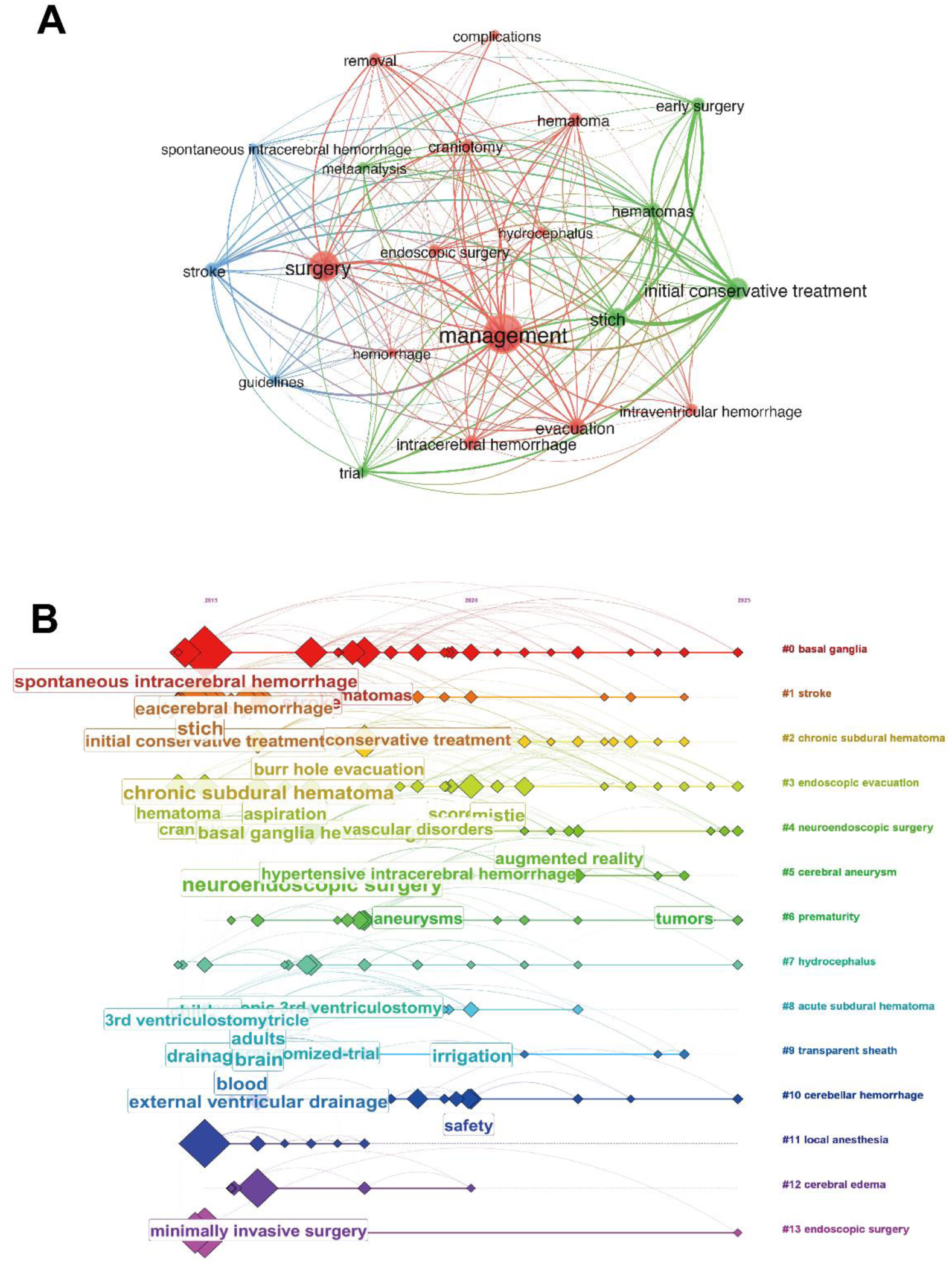
Keywords Plus co-occurrence and temporal evolution. A, Co-occurrence network of Keywords Plus. B, Analysis of Keywords Plus evolution in a timeline view.

The keyword timeline (**Figure 7B**) revealed a two-stage evolutionary pattern. During the early phase (2015–2020), discussions primarily centered on clinical application scenarios, with persistent emphasis on terms such as “spontaneous ICH,” “cerebral hemorrhage,” “chronic subdural hematoma,” “initial conservative treatment,” “external ventricular drainage,” and “aneurysm.” In the later phase (2017–2025), the thematic focus gradually shifted toward technical refinement, exemplified by the emergence of concepts such as “augmented reality.” This temporal transition delineated a trajectory of increasing procedural precision, refined clinical indications, and greater technological integration.

Collectively, research on neuroendoscopic management of ICH converged on a conceptual framework spanning clinical management, surgical intervention, and comparative treatment strategies, evolving from early exploratory applications across heterogeneous contexts toward more defined indications alongside technological advancement.

### 3.7 Source-journal-level contribution and citation structure

Among the 97 journals publishing neuroendoscopic ICH studies, *World Neurosurgery* contributed the largest number of relevant articles (51), followed by *Journal of Neurosurgery* (22) (**Figure 8A**). However, citation impact did not mirror publication volume. *Journal of Neurosurgery* showed the highest mean citation impact among leading journals (27.5 citations per article), followed by *Operative Neurosurgery* (17.6 citations per article). In the citation network (**Figure 8B**), high-frequency journals, particularly *World Neurosurgery* and *Journal of Neurosurgery*, were tightly linked to influential clinical journals such as *Stroke* and *Neurosurgery*, forming a central core. In the co-citation network (**Figure 8C**), *World Neurosurgery* was central within the neurosurgical journal cluster, reflecting its broad thematic scope and integrative role in cross-referencing. Overall, this journal architecture reflected a mature and structured knowledge ecosystem. Core neurosurgical journals stabilized the field’s intellectual foundation, while interconnected specialty journals supported ongoing conceptual expansion and clinical refinement.

**Figure 8.**
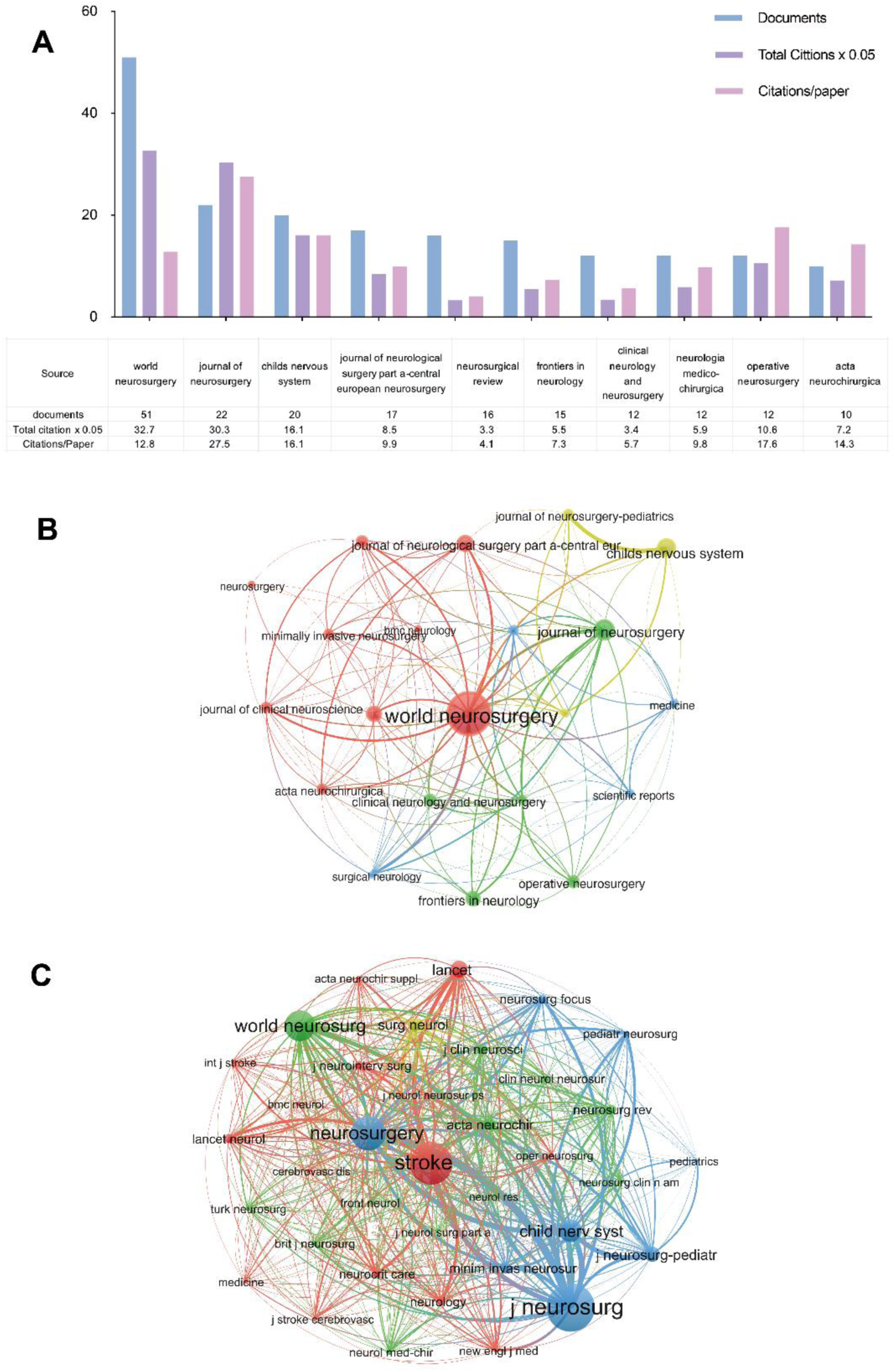
Institutional publishing activity and networks in neuroendoscopic ICH research. A, Top 10 journals with highest publications related to neuroendoscopic ICH. B, The citation network among journals. C, The co-citation network among journals.

### 3.8 Schematic illustration of the six evolutionary phases of ICH therapeutic strategies

This schematic diagram (**Figure 9A-D**) systematically depicts the six sequential therapeutic paradigms for ICH management, including conservative treatment, conventional open craniotomy, small craniotomy, stereotactic hematoma drainage, neuroendoscopic hematoma evacuation, and multimodal comprehensive treatment. Each phase represents a progressive refinement of surgical techniques with the core goal of balancing therapeutic efficacy and iatrogenic injury, reflecting the shift of ICH therapy from traditional invasive intervention to precision-guided minimally invasive and integrated management.

**Figure 9.**
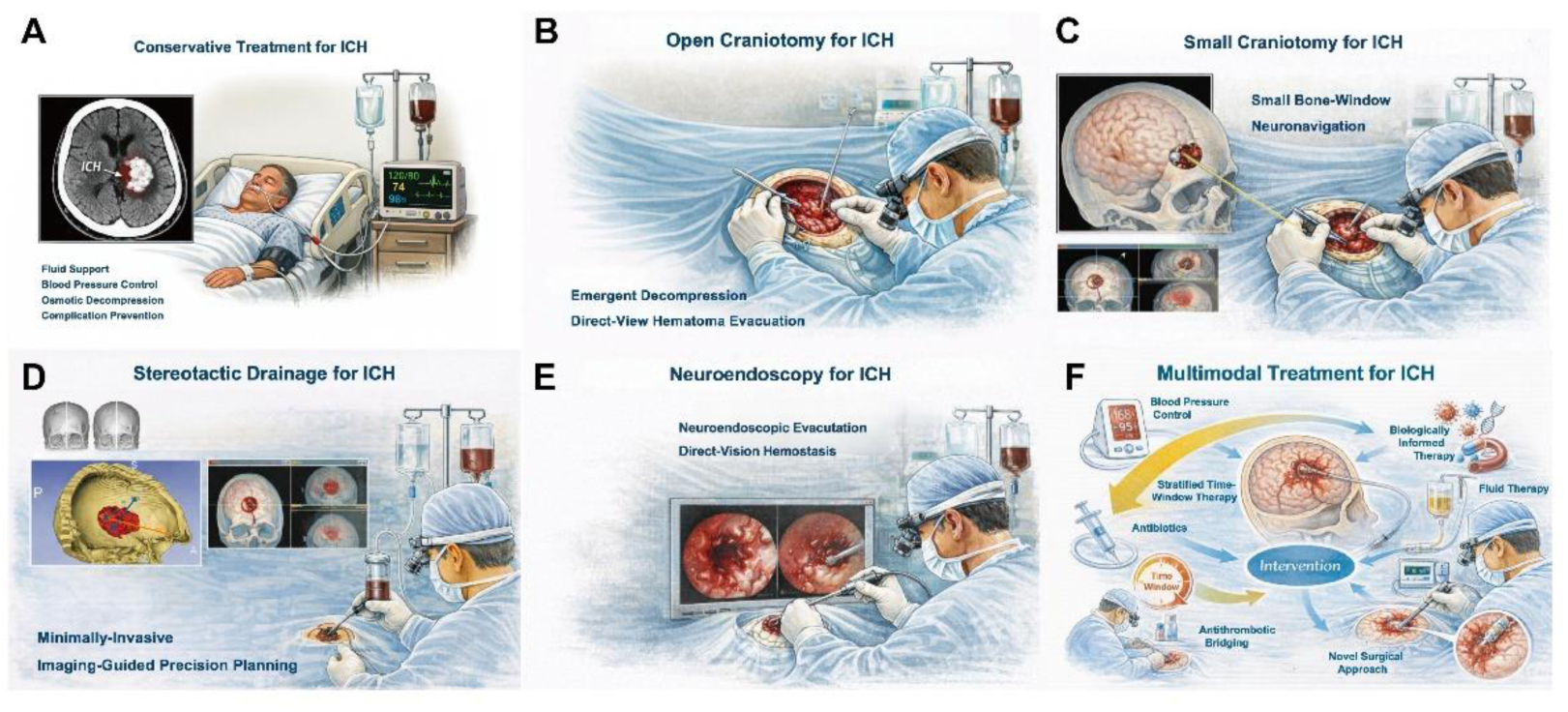
Schematic illustration of the six evolutionary phases of ICH therapeutic. A, Conservative treatment. B, Open craniotomy. C, Small craniotomy. D, Stereotactic drainage. E, neuroendoscopic Hematoma evacuation. F, Multimodal treatment.

## 4 Discussion

This study is the first bibliometric investigation to systematically outline the global research landscape and developmental trajectory of neuroendoscopic treatment for ICH. Scholar attention to this topic rose unevenly, with China and the US co-shaping the field’s core knowledge base. Regionally, North America emphasized evidence-based clinical evaluation, East Asia drove technical and device innovation, and Europe expanded into broader applications, yet cross-border integration remained limited. Clinically, priorities evolved from feasibility and safety to refined patient stratification and long-term effectiveness. The field was shown to transition from an exploratory technical approach toward a more structured, methodologically coherent, and increasingly evidence-oriented research domain.

The ICH therapeutic paradigm progressed through six phases—conservative care, conventional craniotomy, small craniotomy, stereotactic aspiration, neuroendoscopy, and multimodal strategies—with stepwise technical refinement and escalating evidence-based scrutiny.

The first phase of ICH management was characterized by a predominant reliance on conservative medical management [41]. Therapeutic strategies primarily focused on blood pressure control, intracranial pressure control, infection prevention, and neurocritical care support to minimize secondary neurological deterioration [42]. Conservative treatment built a “stabilization-oriented” paradigm, implicitly defining the concept that patient outcomes were shaped not only by the primary hemorrhagic insult itself, but also by the subsequent cascade of physiological and metabolic disturbances [43]. This paradigm was a conceptual framework that later re-emerged in more mechanistic and pathophysiological models of ICH progression.

In the second phase, as neuroimaging and operative techniques advanced, open craniotomy was regarded as the most straightforward surgical strategy. However, subsequent high-quality RCT evidence progressively tempered this early confidence. In comparison with conservative management, The STICH trial [8] showed that early surgery did not improve favorable 6-month outcomes (p = 0.414), and STICH II [9] similarly demonstrated no significant benefit in superficial lobar ICH (p = 0.367). More recently, the SWITCH trial [10] in severe deep ICH (basal ganglia/thalamus) showed borderline 180-day mRS 5–6 reduction (p = 0.057). A meta-analysis by Wilting et al. [44] pooling RCTs found conventional craniotomy may improve functional outcomes but with substantial uncertainty (risk ratio 1.41, 95% CI 0.77 to 2.55). Taken together, these landmark trials reframed craniotomy in ICH by highlighting surgical injury and patient selection as key constraints, redirecting efforts toward technique refinement to reduce iatrogenic harm and optimize patient stratification.

In the third stage, small bone window craniotomy was developed in response to pessimistic evidence on conventional craniotomy. Burr hole drainage was increasingly adopted and associated with shorter procedure times, reduced hospital stays, earlier clinical improvement, and more favorable recovery of daily functional capacity [45]. Small bone window craniotomy reduced surgery-related injury, and reflected a shift toward tissue-preserving surgical philosophies.

The fourth stage advanced surgical minimalism through stereotactic and catheter-based techniques. A meta-analysis by Scaggiante et al. [11] reported that stereotactic thrombolysis was associated with significantly lower death or dependence (OR 0.47, 95% CI 0.34–0.65) compared with conventional treatments. However, in the SICHPA trial [46], 180-day mortality did not differ between stereotactic catheter drainage and conservative therapy (p = 0.84). Similarly, the MISTIE II trial [47] reported no significant difference in 180-day functional independence (mRS ≤ 3) between medical management and the catheter-based evacuation plus rt-PA (p = 0.26). at 365 days, MISTIE III [31] likewise showed no significant increase in functional independence, despite fewer 30-day serious adverse events (p = 0.012). Collectively, these findings suggest that procedural minimalism did not guarantee improved long-term outcomes, highlighting the need for context-specific, high-quality evidence.

As the fifth-stage strategy, neuroendoscopic evacuation integrated prior approaches while mitigating their structural limitations. Neuroendoscopy uses a smaller corridor with less parenchymal injury than craniotomy [48] and allows direct visualization with more effective evacuation and hemostasis than stereotactic catheter drainage [49]. Accordingly, a meta-analysis of RCTs by Scaggiante et al. [11] suggested that endoscopic techniques demonstrated advantages over conventional strategies in supratentorial ICH. Gui et al. [50], in a retrospective cohort study, found neuroendoscopy achieved shorter operative time, less blood loss, and higher evacuation rates than small bone window craniotomy (all p < 0.05). A network meta-analysis of 26 RCTs by Wang et al. [51] reported lower mortality versus standard medical care (RR 0.66, 95% CI 0.50–0.87). These findings positioned neuroendoscopy as a distinct surgical paradigm within the MIS continuum.

In the sixth paradigm, surgery was positioned within a multimodal framework informed by secondary injury biology, including neuroinflammation (e.g. microglia and neutrophil extracellular traps [52]), blood–brain barrier disruption [53], iron-mediated cytotoxicity [54], and neurovascular unit dysfunction [55]. Moreover, parallel technological advances reshaped neuroendoscopic workflows. Wu et al. [56] reported that robot-assisted neuroendoscopy with ICP monitoring reduced intraoperative blood loss (p = 0.001) and improved discharge mRS (p = 0.011) versus conventional neuroendoscopy. Advances in robotic stereotaxy, navigation, and workflow standardization further signal a move toward reproducible, precision-guided paradigms [57,58].

Overall, ICH therapy evolved from stabilization-focused care to precision-guided intervention. Neuroendoscopy became pivotal, and future progress will hinge on multimodal, stratified management.

## 5 Limitations

This study has several limitations. First, we only included articles indexed in the WOSCC, potentially missing records from other databases. However, WOSCC offers standardized cited-reference data that ensures reproducibility and reduces cross-database noise. Second, we limited inclusion to English publications. Nonetheless, this restriction strengthens metadata consistency, enabling more reliable disambiguation and keyword clustering.

## 6 Conclusion

This study provided the first bibliometric mapping of the global research landscape of neuroendoscopic intervention for ICH. Overall, the field has progressed from establishing procedural safety and feasibility, to conducting evidence-generating clinical trials, and further to refining indications, alongside continual technological advances. Looking forward, priorities include sustained technical innovation, strengthening high-level clinical evidence, and further defining the indications for neuroendoscopy.

## Data Availability

No new data were generated or analysed in this study.

## Abbreviations

ICH: Intracranial Hemorrhage;
MIS: Minimally Invasive Surgery;
IVH: Intraventricular hemorrhage;
SAH: Subarachnoid hemorrhage;
OR: Odds ratio;
CI: Confidence interval;
RCTs: Randomized controlled trials;
WOSCC: Web of Science Core Collection;
ICP: Intracranial pressure;
rt-PA: Recombinant tissue plasminogen activator;
mRS: Modified Rankin Scale;
ICU: Intensive Care Unit;
STICH: Surgical Trial in Intracerebral Haemorrhage;
SWITCH: Decompressive Craniectomy for Severe Deep Intracerebral Hemorrhage Trial;
MISTIE: Minimally Invasive Surgery with Thrombolysis in Intracerebral Haemorrhage Evacuation;
SICHPA: Stereotactic Intracerebral Hematoma Plasminogen Activator trial;
INET: Intra-neuroendoscopic technique;
NESICH: Neuroendoscopic parafascicular evacuation of spontaneous intracerebral hemorrhage

## Acknowledgements

We express our gratitude to all researchers and patients who have contributed significantly to advancing the field of neuroendoscopic ICH.

